# The Long Road to Care: A Mixed Methods Analysis of Referral Delays for Pediatric Injury Patients in Northern Tanzania

**DOI:** 10.64898/2026.10.06.26364512

**Authors:** Elizabeth M. Keating, Rosalia Njau, Joaquim Magoma, Baraka Moshi, Deusdedith Marandu, Emmanuel Minja, Itikija Msuya, Anord Anthony, Francis Sakita, Matti Koenig, Edwin Shewiyo, Cassidy Huun, Sharla Rent, Getrude Nkini, Nancy Mmary, Heather T. Keenan, Jennie L. Hill, Melissa H. Watt, Catherine A. Staton, Blandina Theophil Mmbaga, Joao Ricardo Nickenig Vissoci

## Abstract

Every day, 1900 children die from an injury, with the majority in low-and middle-income countries. In Tanzania, reaching definitive care, or the appropriate final level of care for a child’s injury, often requires referral to a higher level facility. This hierarchical system introduces challenges that may lead to delayed care and, for severely injured children, cause mortality. The aim was to identify patient, healthcare provider, and health system-level barriers to timely referral for pediatric injury patients at the first site of care in order to identify targets for an intervention to reduce time to definitive care. In this mixed-methods study, a pediatric injury registry was established at three health facilities spanning three health system levels in Northern Tanzania. Thirty family caregivers of pediatric injury patients participated in interviews, while healthcare providers participated in four focus group discussions. The Consolidated Framework for Implementation Research framed interview guides and thematic analysis of the qualitative data. Of 782 patients enrolled in the pediatric injury registry from Aug 2024-Aug 2025, 269 were referred. Most visited 1-4 facilities before reaching definitive care (range 1-6). The median time from injury to definitive care was 15.1 hours (IQR 1.8-33). Qualitative data collection with family caregivers and healthcare providers revealed barriers to explain the delays to care in the outer setting (such as non-standardized referral pathways), the inner setting (such as delays in the decision to refer), and individual characteristics (such as inadequate training in pediatric injury care). According to the World Health Organization standards, every child who needs referral should be sent within 2 hours of arrival. To improve the referral delays found in this study, a locally co-designed, multicomponent health systems intervention for healthcare providers at the first site of care must be developed that includes pediatric injury care training and standardized referral decision support.

**Key Messages:**

- Injuries are important contributors to child morbidity and mortality globally, with low-and middle-income settings like Tanzania disproportionately affected.
- In Tanzania there is a complex health system where pediatric injury patients present to primary health facilities first, and are referred up the health system until they reach the definitive care that they need.
- Barriers at the outer setting, inner setting, and individual level of the health system contribute to delays in reaching definitive care.
- In order to combat these delays and improve child morbidity and mortality, a locally co-designed, multicomponent health systems intervention at the level of the first site of care must be developed that includes standardized referral decision processes.

## Introduction

Every day, 1900 children die from an injury and more than 95% of these deaths occur in low-and-middle-income countries (LMICs).(2008; Abdur-Rahman et al. 2012; Collaborators 2018) The proportion of those affected is even larger in Sub-Saharan Africa.(Abdur-Rahman et al. 2012) Thus, the burden of pediatric injuries in LMICs is a significant public health problem that deserves urgent attention.(2008; St-Louis et al. 2020) Yet, there is a lack of data on strategies to improve the care and outcomes of injured children in LMICs.(Keating et al. 2019; WHO) Most research on pediatric injuries in LMICs uses registry data from tertiary hospitals to assess the epidemiology of injuries (Bradshaw et al. 2018; Burstein et al. 2016; Petroze et al. 2020; St-Louis et al. 2020; Traynor et al. 2020) but does not examine how injured children reach tertiary care from the site of injury.

Pediatric injury morbidity and mortality can be improved with prompt arrival to definitive care, defined as the appropriate level of final care for the injury severity and type. Research suggests that delaying arrival to definitive care in children is associated with higher mortality risk.(Botchey et al. 2017; Boyd & Cowley 1983; Jin et al. 2021; Mock et al. 2009; Rogers et al. 2015; Sa’avu et al. 2014; Wisborg et al. 2011) In Northern Tanzania, the public health system is organized to conserve resources and facilitate progressive access to more specialized services.(Aghan et al. 2023; Nicks et al. 2012) Three levels of increasingly specialized care are common: the primary level (health center or district hospital), the secondary level (regional hospital), and the tertiary level (zonal referral hospital). In preliminary work at the tertiary zonal referral hospital Kilimanjaro Christian Medical Centre (KCMC) in Northern Tanzania, 81% of injured children presented to at least one primary health facility before referral to definitive care.(Keating et al. 2022a) Delays in reaching KCMC were identified, including a complex referral system that requires families to seek care at a primary health facility before being referred stepwise to higher levels of care.(Keating et al. 2022a) This hierarchical system of care introduces logistical challenges that may lead to delayed care and, for severely injured children or those needing specialist care, cause unnecessary and sometimes deadly delays.

Thus, there is an opportunity to streamline the referral system by implementing an intervention at the primary health facility, the first site of care, to bring injured children to definitive care more quickly. Expediting care within the healthcare system requires an intervention rooted in implementation science, as this allows for examination of factors of the inner setting, outer setting, and individual level that influence care and could be targeted with such an intervention. Implementation science frameworks, such as the Consolidated Framework for Implementation Research (CFIR), are useful for developing or adapting interventions to improve care and outcomes. CFIR was chosen for this study as it has been successfully used in many similar LMIC settings,(Brown et al. 2021; Kumar et al. 2020; Le et al. 2022; Means et al. 2020b; Michaud-Létourneau et al. 2022; Ojo et al. 2021; Peiffer-Smadja et al. 2020; Romani et al. 2020; Taylor Salisbury et al. 2021; Thomas et al. 2022; Turner et al. 2021), has been used by this authorship team,(Joiner et al. 2023; Sanuade et al. 2025) and aligns closely with the need to understand multilevel health systems barriers in order to inform a systematic assessment of multilevel implementation contexts.(Damschroder et al. 2022)

To inform the development of a health system intervention at the first site of care aimed at reducing time to definitive care, a CFIR-guided mixed-methods study of pediatric injury care at two common first sites of care was conducted. The study aim was to identify patient-, provider-, and systems-level barriers to timely definitive care for pediatric injury patients starting at the first site of care to inform the development of an intervention to reduce time to definitive care in Northern Tanzania.

## Methods

### Study design

This explanatory mixed-methods study used three data sources: quantitative registry data, qualitative semi-structured in-depth interviews (IDIs) with family caregivers, and focus group discussions (FGDs) with healthcare providers to both identify delays and explore their underlying causes. An injury registry was first created spanning the healthcare system at two common first sites of care for pediatric injury patients in Northern Tanzania, Pasua Health Centre and Hai District Hospital, and a tertiary referral center, KCMC (Figure 1). This quantitative registry data was used to describe referral patterns and time to each higher level of care in pediatric injury patients. Patients were defined as being referred if they were sent from one level of care to a higher level of care and thus visited more than one health facility.

**Figure 1.**
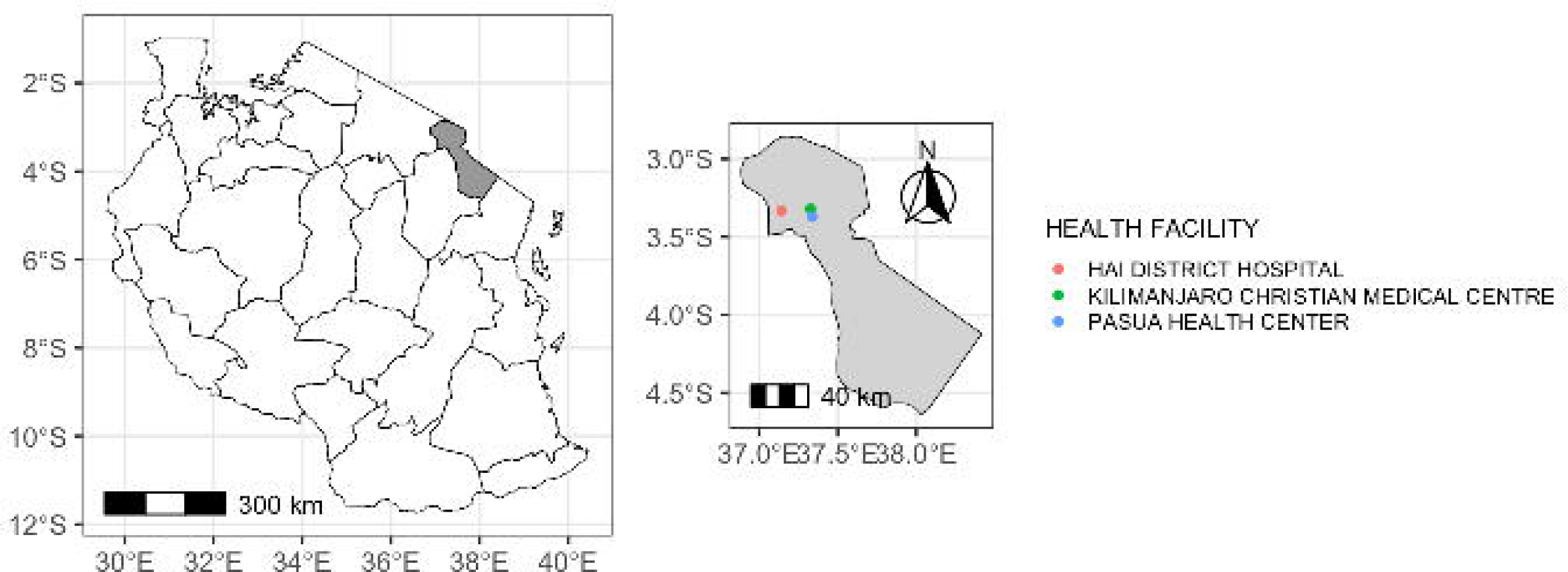
Map of Tanzania with the Kilimanjaro region in the inset, showing the three health facilities where data collection was completed. Alt text: Map of Tanzania showing the three health facilities where data collection was completed.

Definitive care was defined as the final health facility visited by each patient, regardless of the level of the health system. Using qualitative IDIs and FGDs with family caregivers and healthcare providers, barriers to prompt referrals were explored. The quantitative and qualitative data were mapped to the CFIR determinants of outer setting, inner setting, and individual characteristics.(Damschroder et al. 2022) This study received ethical approval from the institutional review boards at the Tanzania National Institute for Medical Research (approved 5/21/2024), and the authors’ institutions (approved 1/22/2024 and 7/27/2023).

### Study setting

The study took place within the KCMC referral system at three health facilities in Northern Tanzania. In addition to KCMC, two health facilities that are common first sites of care for injured children were included: Pasua Health Centre and Hai District Hospital. These sites were selected for proximity, logistical ease, representativeness, and the number of injured children they refer to KCMC. The two sites differ in the level of available care and distance from KCMC, improving the generalizability of the results. Pasua Health Centre is a government health center in urban Moshi, Tanzania, approximately 20 minutes by car from KCMC. Hai District Hospital is a government district hospital in semi-urban Hai, Tanzania, approximately 35 minutes by car from KCMC. Both Pasua Health Centre and Hai District Hospital have an Outpatient Department (OPD) and a pediatric ward where they receive and admit injured children. Both have access to basic first aid, laboratory examinations, and X-ray and ultrasound (though no radiologist to interpret images). Hai District Hospital has an orthopedic surgeon, and Pasua Health Centre has a general surgeon; otherwise, there are no specialists at either facility, and most care is provided by medical officers, who are medical doctors with basic training. KCMC is one of four zonal referral hospitals in Tanzania, serving the northern regions. KCMC has a catchment population of 12 million people and is in the Kilimanjaro region of northern Tanzania. It serves as a referral site for children with injuries that require specialized investigations, imaging, surgery, and treatment. Specialized services available include pediatricians, pediatric surgeons, orthopedic surgeons, neurosurgeons, ophthalmologists, and a specialized burn center.

### Quantitative Registry Data

#### Participants

The KCMC pediatric injury registry was launched in November 2020 to identify quality improvement opportunities for the care of pediatric trauma patients.(Keating et al. 2022a; Keating et al. 2022b) This registry was expanded to include Pasua Health Centre and Hai District Hospital in August 2024. Patients were enrolled in the registry on presentation to the KCMC Emergency Medical Department (EMD), Pasua Health Centre OPD, or Hai District Hospital OPD if they were less than 18 years old and presented with an injury sustained in the past 6 months. The registry enrolled patients who were injured with injury types including burns, fractures, lacerations, traumatic brain injuries, ingestions/poisonings, animal envenomation, road traffic injuries, falls, drownings, penetrating trauma, non-accidental trauma, and rape. Patients who presented for injury follow-up care were excluded. Patients were asked to recall their journey to get to the health facility where they were enrolled at the time of presentation and registry enrollment. If a patient was enrolled in the registry at Hai District Hospital or Pasua Health Centre, and then referred to KCMC, their data collection was continued on the same patient ID at both facilities. Thus, these patients were followed prospectively through the health system.

#### Procedures

The pediatric injury registry prospectively enrolled all patients. All registry data were recorded on tablets in REDCap,(Harris et al. 2009) and the quality of all entries was reviewed by the principal investigator (EMK). The following REDCap database variables were extracted for this analysis: demographics, mechanism of injury, time of injury, mechanism of transport, each health facility that provided treatment, time of arrival to each health facility in the care pathway, and patient outcomes, including in-hospital mortality and morbidity. Morbidity was measured using the Glasgow Outcome Score-Extended Pediatrics (GOS-E Peds). The GOS-E Peds is an eight item instrument designed to assess outcomes in children after traumatic brain injury and has been expanded for use in the overall injured child population. (Ardolino et al. 2012; Beers et al. 2012; Williamson et al. 2011) During registry data collection, research assistants scored the GOS-E Peds by asking the family caregiver about the child’s level of functioning across multiple domains. As others have done previously, the score was dichotomized, with 1-2 representing “good recovery” and 3-8 representing “poor recovery”.(Gabbe et al. 2014; Keating et al. 2022b) Patients with missing outcomes were excluded.

### Qualitative Interviews with Family Caregivers

#### Participants

Semi-structured IDIs were completed with 30 family caregivers of injured pediatric patients. Nine family caregivers were male and 21 were female, and ages of participants ranged from 24 to 77 years. Family caregivers of injured children were recruited by a research assistant at one of three health facilities for a semi-structured IDI if they were the caregiver (such as mother, father, or other relative caring for the child) of an injured child who was initially seen at Pasua Health Center or Hai District Hospital and referred to a higher level of care. Other inclusion criteria included fluency in Swahili and the ability to provide informed consent. Participants were provided a copy of the consent form, which was read aloud by research assistants. Participants provided written informed consent prior to completing the qualitative interview; those unable to write provided a thumbprint in the presence of a witness. Participants received 10,000 Tanzanian shillings (approximately 4 U.S. dollars) for their time and transport. The number of participants to be recruited was preset at 30 and informed by team-based discussions of when thematic saturation was expected to occur.(Guest et al. 2006)

#### Procedures

The interview guide was informed by the CFIR 2.0 to identify health systems-level barriers and challenges in the care of pediatric injury patients across the care continuum, from injury occurrence through presentation at the health facility to disposition and referral. Family caregiver participants were asked: how the injury occurred, when the decision was made to seek care, where they went to receive care, why they chose to go there first, the care received at the health facility, why they were referred to a higher level of care, facilitators and barriers in the process, and what would have been helpful in the care of the child. The guide was translated from English into Swahili by three native speakers, checked for accuracy in translation meetings by a team of native speakers, and pilot-tested by the research team for comprehension and question flow.

Interviews were conducted between August 2024 and July 2025. All interviews were completed in Swahili by three bilingual, trained Tanzanian research assistants with extensive experience in qualitative interviewing. The research assistants completed two weeks of study-specific training, including review of the study protocol and qualitative interview guides. IDIs were conducted in quiet, private locations within the study facilities. In some cases, when the patient had already been discharged home before the research assistant learned of the admission, participants were consented, and interviews were conducted over the phone. Each interview took approximately one hour. Interviews were audio-recorded with participants’ consent. Interview audio recordings were simultaneously transcribed and translated to English by the three bilingual Tanzanian research assistants.

### Qualitative Focus Group Discussions with Healthcare Providers

#### Participants

Four FGDs were completed with 31 healthcare providers who provide care for injured pediatric patients at Pasua Health Center and Hai District Hospital. Seven healthcare provider participants were male and 24 were female, and ages of participants ranged from 25 to 58 years. Participants included 10 medical doctors, 3 clinical officers, 17 nurses, and 1 physiotherapist, and the length of experience in their position ranged from 1 year to 28 years. Healthcare providers were eligible to participate in an FGD if they worked in the outpatient department or the pediatric ward at each facility. These departments were selected because they care for pediatric injury patients at these facilities. Our FGD participants were fluent in Swahili and were able to provide informed consent. Participants were provided a copy of the consent form, which was read aloud by research assistants. Participants provided their written informed consent prior to completing the qualitative interview. Healthcare providers were selected for recruitment by the medical officer in charge of each facility. After the medical officer provided our research assistant with the list of names, individuals were recruited by a research assistant face to face and scheduled to participate in a FGD. To adequately represent healthcare providers caring for the pediatric injury population, healthcare providers with a variety of specialties, experience, and ages were interviewed. Participants received 20,000 Tanzanian shillings (approximately 8 U.S. dollars) for their time and transport. The number of FGDs was preset to four and informed by team-based discussions of when thematic saturation was likely to occur.

#### Procedures

The CFIR 2.0 was used to frame the FGD and was aligned with the aim of understanding systems-level barriers and challenges in the care of pediatric injury patients throughout the care continuum, from injury occurrence to presentation to disposition (admission to the ward, transfer, or discharge home). Healthcare providers were asked about: pediatric injury patient pre-hospital care, patient assessment, patient stabilization, patient disposition, patient referral, and intervention ideas to improve care for pediatric injury patients. The guide was translated from English into Swahili by three native speakers, checked for accuracy in translation meetings by a team of native speakers, and pilot-tested by the research team. FGDs were conducted between August 2024 and April 2025. All FGDs were conducted in Swahili by three trained Tanzanian research assistants with extensive experience in qualitative interviewing. The research assistants completed two weeks of training, including qualitative interviewing strategies and familiarity with the study protocol and qualitative interview guides. FGDs were conducted in quiet, private locations within the study facilities. Each FGD took approximately 2 hours. Interviews were audio recorded for verbatim transcription. Interview audio recordings were transcribed and translated to English by the three bilingual Tanzanian research assistants.

### Quantitative data analysis

To characterize health systems’ delays of injured children, descriptive statistics summarized demographics, timing of care at each level, clinical characteristics, and referral pathways. Continuous data were represented as means with standard deviation or medians with interquartile ranges. Time was defined as the median time from injury to reaching definitive care in hours.

Kaplan-Meier curves were built to depict time to definitive care comparing the number of hospitals each participant went to. Patients with good versus poor outcomes were compared across demographic and treatment characteristics using Fisher’s exact test or Chi-Square tests, as appropriate. Patients that were referred and not referred from the first sites of care were compared using Fisher’s exact test, or Chi-squared tests as appropriate. Cox proportional hazards regression was performed to identify factors associated with shorter time to definitive care.

Covariates included age group, sex, residency, living arrangement, ambulance use, number of hospitals visited, and insurance status. Hazard ratios with 95% confidence intervals were reported. All analyses were performed in STATA (version 18.0) and figures drawn using R (version 4.4.1).

### Qualitative data analysis

The theory-informed IDI and FGD guides served as a deductive framework for developing the preliminary codebook. After reviewing the transcripts, the research team met to reach consensus on the emerging themes and create a codebook organized around the CFIR framework domains. The codebook was then uploaded into Dedoose© qualitative coding software, and the transcripts were double-coded in Dedoose© by two members of the coding team. The first coder performed the intial qualitative coding, and the second coder reviewed the coding and made notes anywhere there were discrepancies. Intercoder discrepancies were discussed among the research team during coding meetings and resolved by consensus, with EMK determining the final code as needed. The codebook was continually adapted to reflect new or emerging themes as coding progressed. The coding team included members from Tanzania for a cultural insider perspective and from the US for a cultural outsider perspective. IDIs and FGDs were analyzed using the CFIR framework to identify barriers at the inner and outer setting levels and characteristics of individuals.(Damschroder et al. 2022; Safaeinili et al. 2020)

## Results

782 patients were enrolled in the pediatric injury registry at the three study sites within the KCMC catchment area. A total of 269 patients were seen at an initial health facility and had either been referred in, or were referred out for definitive care. The median age was 6 years (IQR 4,10), and 32.5% were female. Falls, blunt force, burns, and road traffic injuries were the most common injury mechanisms (Table 1). Overall mortality was 3.6% (n=10). The demographic and clinical characteristics of participants differed significantly between those referred and those not referred, including enrollment location, age group, residency, living arrangement, mechanism of injury, payment method, number of health facilities visited, and outcome measured by the GOS-E Peds (Table 1).

**Table 1.** Demographic and clinical characteristics of patients comparing those who were referred from one health facility to another (either in or out), and non-referrals.

| Variable | Total<br>N (%) | Referrals<br>N (%) | Non-Referrals<br>N (%) | P-value |
| --- | --- | --- | --- | --- |
| **Age Group** |  |  |  | 0.002* |
| Infant (0-1 year) | 57 (7.5) | 13 (4.9) | 44 (8.9) |  |
| Toddler (2-3 years) | 119 (15.7) | 42 (15.9) | 77 (15.6) |  |
| Preschool (4-5 years) | 134 (17.7) | 60 (22.8) | 74 (15.0) |  |
| Child (6-11 years) | 295 (39.0) | 84 (31.9) | 211 (42.7) |  |
| Preteen (12-14 years) | 80 (10.6) | 35 (13.3) | 45 (9.1) |  |
| Teen (15-17 years) | 72 (9.5) | 29 (11.0) | 43 (8.7) |  |
| **Sex** |  |  |  | 0.738 |
| Male | 528 (67.5) | 187 (68.3) | 341 (67.1) |  |
| Female | 254 (32.5) | 87 (31.7) | 167 (32.9) |  |
| **Lives With** |  |  |  | <0.001* |
| Single parent | 69 (8.8) | 36 (13.2) | 33 (6.5) |  |
| Both parents | 657 (84.1) | 209 (76.6) | 448 (88.2) |  |
| Grandparent or Other | 55 (7.0) | 28 (10.3) | 27 (5.3) |  |
| **Number of children living in the home, N (%)** |  |  |  | 0.001* |
| 1 (patient) | 74 (9.5) | 32 (11.7) | 42 (8.3) |  |
| 2-3 | 484 (61.9) | 150 (54.7) | 334 (65.7) |  |
| 4-5 | 193 (24.7) | 73 (26.6) | 120 (23.6) |  |
| >=6 | 31 (4.0) | 19 (6.9) | 12 (2.4) |  |
| **Mechanism of Injury** |  |  |  | <0.001* |
| Road Traffic Injury | 102 (13.1) | 72 (26.4) | 30 (5.9) |  |
| Blunt force/Stuck/Hit | 136 (17.5) | 77 (28.2) | 59 (11.7) |  |
| Fall | 376 (48.3) | 72 (26.4) | 304 (60.2) |  |
| Burn | 118 (15.2) | 42 (15.4) | 76 (15.1) |  |
| Other | 46 (5.9) | 10 (3.7) | 36 (7.1) |  |
| **Payment method** |  |  |  | <0.001* |
| Personal/family cash | 684 (87.5) | 227 (82.6) | 457 (89.9) |  |
| Insurance | 70(8.9) | 37 (13.5) | 33 (6.5) |  |
| Hospital exemption or child under 5 | 22 (2.8) | 4 (1.5) | 18 (3.5) |  |
| None and abscond | 6 (0.8) | 6 (2.2) | 0 (0.0) |  |
| **Total Number of Health Facilities Visited** |  |  |  | <0.001* |
| 1 | 507 (64.8) | 0 (0.0) | 507 (99.8) |  |
| 2 | 151 (19.3) | 151 (55.1) | 0 (0.0) |  |
| 3 | 107 (13.7) | 106 (38.7) | 0 (0,0) |  |
| 4 to 6 | 17 (2.2) | 17 (6.2) | 0 (0.0) |  |
| **GOS-E Peds** |  |  |  | <0.001* |
| Good outcome (1-2) | 712 (92.7) | 225 (84.6) | 487 (97.0) |  |
| Poor outcome (3-8) | 56 (7.3) | 41 (15.4) | 15 (3.0) |  |

**\*Mechanism of Injury Other** = Penetrating traumas, drowning, suffocation/choking/hanging, Poisoning/toxic exposure, animal envenomation, child abuse.

Definitive care was defined as the final health facility, and could be any level of the health system. Most (97.4%) patients visited between one and three health facilities before reaching definitive care (range 1-6). Patients visiting only one health facility were non-referrals. The median time from injury to the final (definitive) facility was 15.1 hours (IQR 1.8-33) for all patients, regardless of the last facility they visited. The median time from injury to the first facility was 2.1 hours (IQR 0.7–18.1). Among inter-facility referrals, the median time from the first to the second facility was 7.6 hours (IQR 2.7–30.0), from the second to the third 21.3 hours (IQR 4.7–52.5), and from the third to the fourth 24.2 hours (IQR 8.0–40.3) (Figure 2). Overall, the time from injury to each subsequent facility increased progressively across the referral chain (Figure 3).

**Figure 2.**
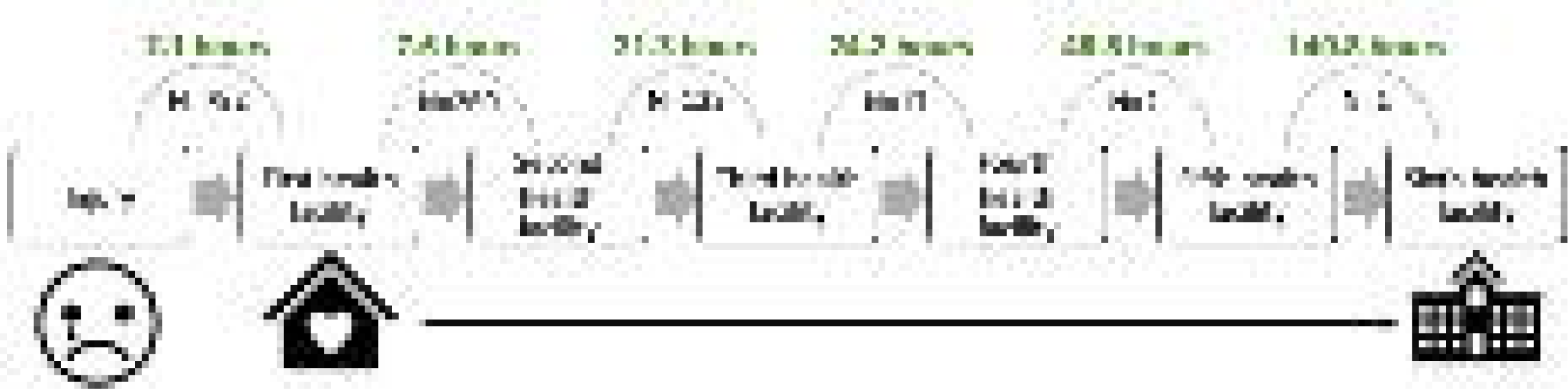
Pathway to care for pediatric injury patients in our study showing median times at each health system segment.* *KCMC could be anywhere from the first to the sixth health facility. Alt text: Figure showing the pathway to care for pediatric injury patients in our study showing median times for each health system segment, from the injury to the first health facility to the sixth health facility.

**Figure 3.**
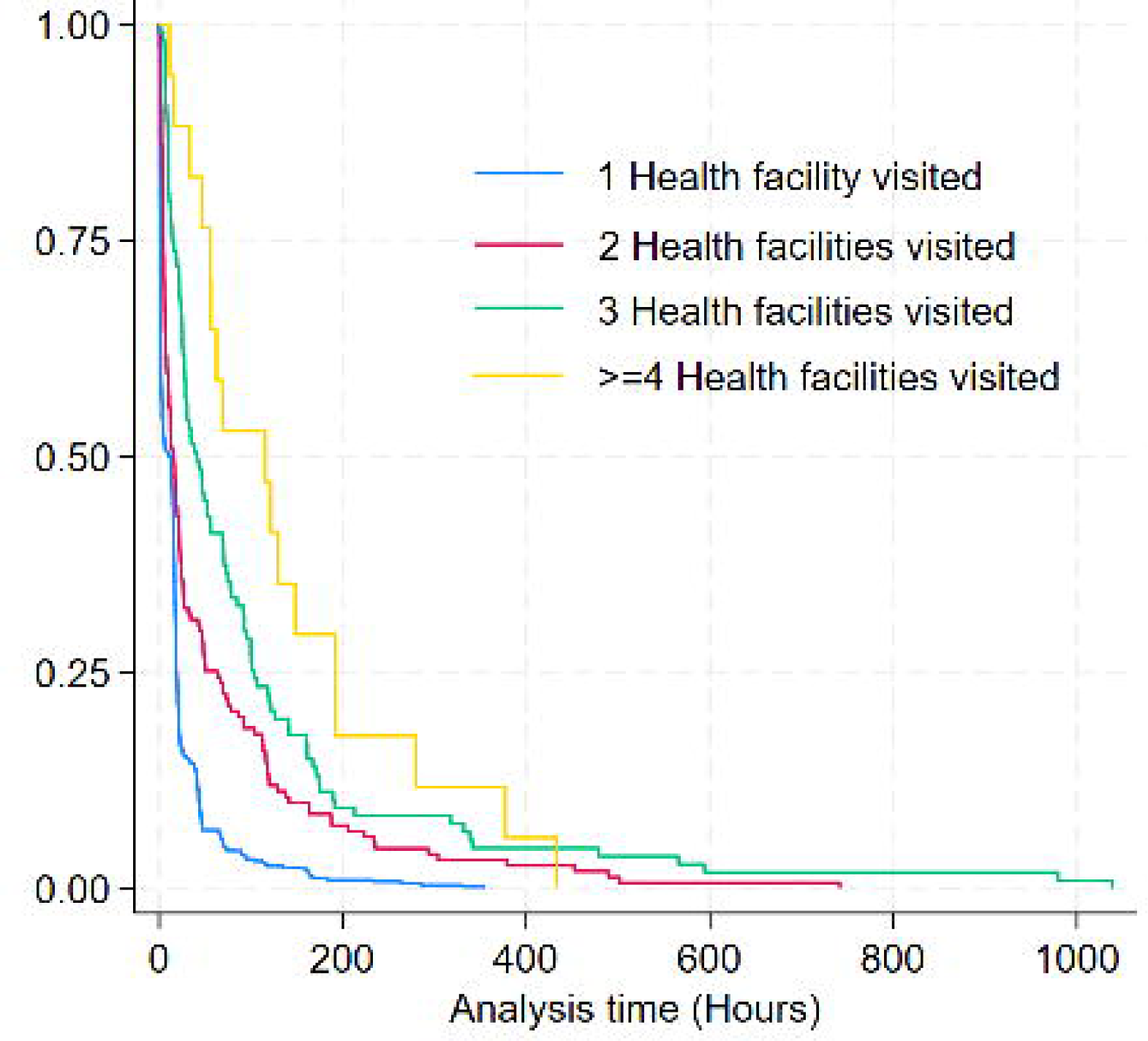
Kaplan-Meier survival curves showing time to definitive care among patients referred from 1 to 6 health facilities before reaching definitive care. Alt text: Graph depicting Kaplan-Meier survival curves showing time to definitive care among patients referred from 1 to 6 health facilities before reaching definitive care.

In the adjusted Cox proportional hazards regression, factors significantly associated with faster time to definitive care include being an infant (0-1 year old), residing in Hai District, having a road traffic injury as the mechanism of injury, being transported by ambulance, having insurance, and requiring fewer referrals to health facilities (Table 2). Factors significantly associated with delayed time to definitive care include being a child older than 1 year, residing in rural Moshi compared with urban Moshi, living with a grandparent or other caregiver who is not a parent, having a mechanism of injury other than road traffic injury, and visiting a greater number of health facilities.

**Table 2.**
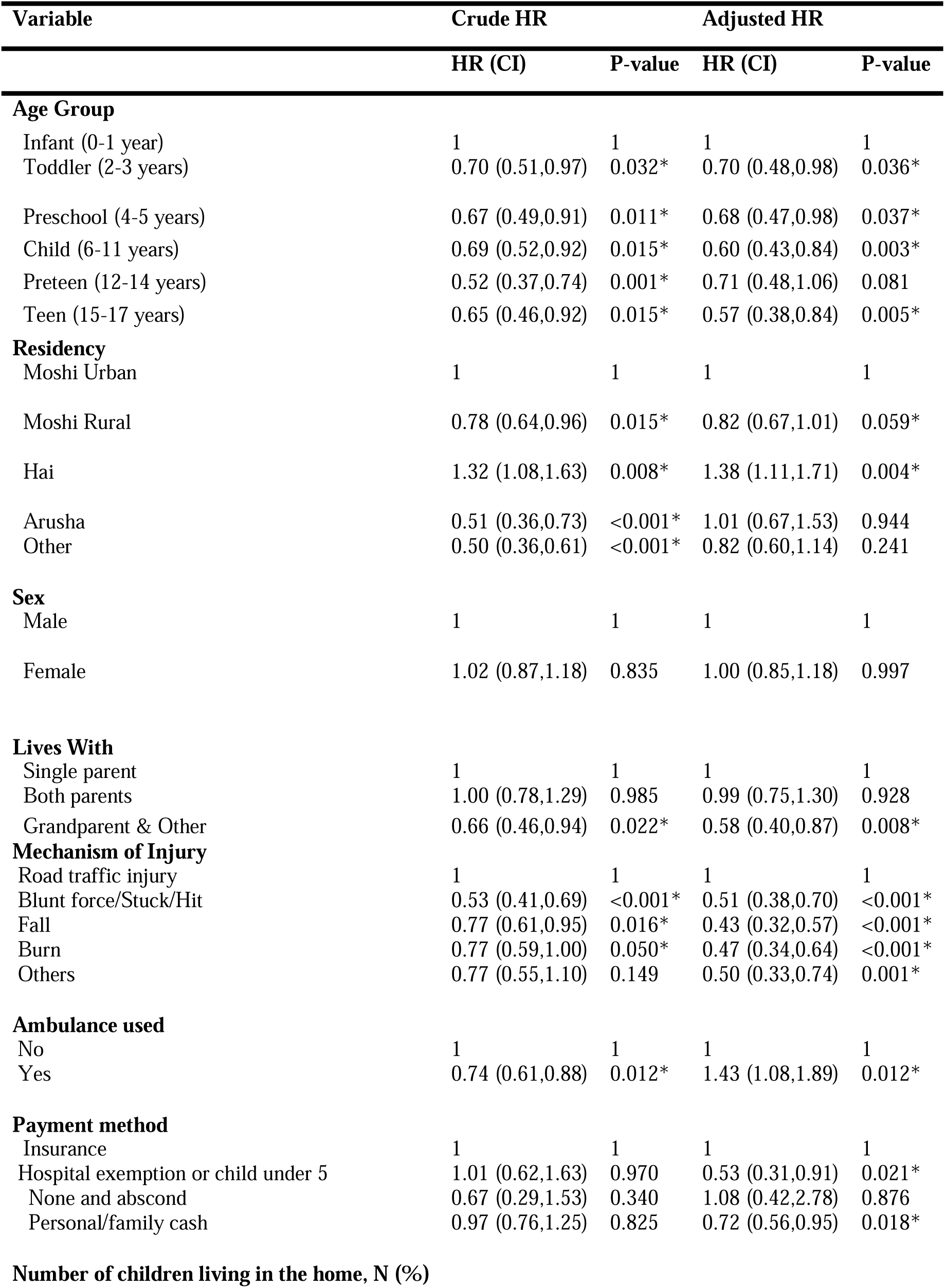
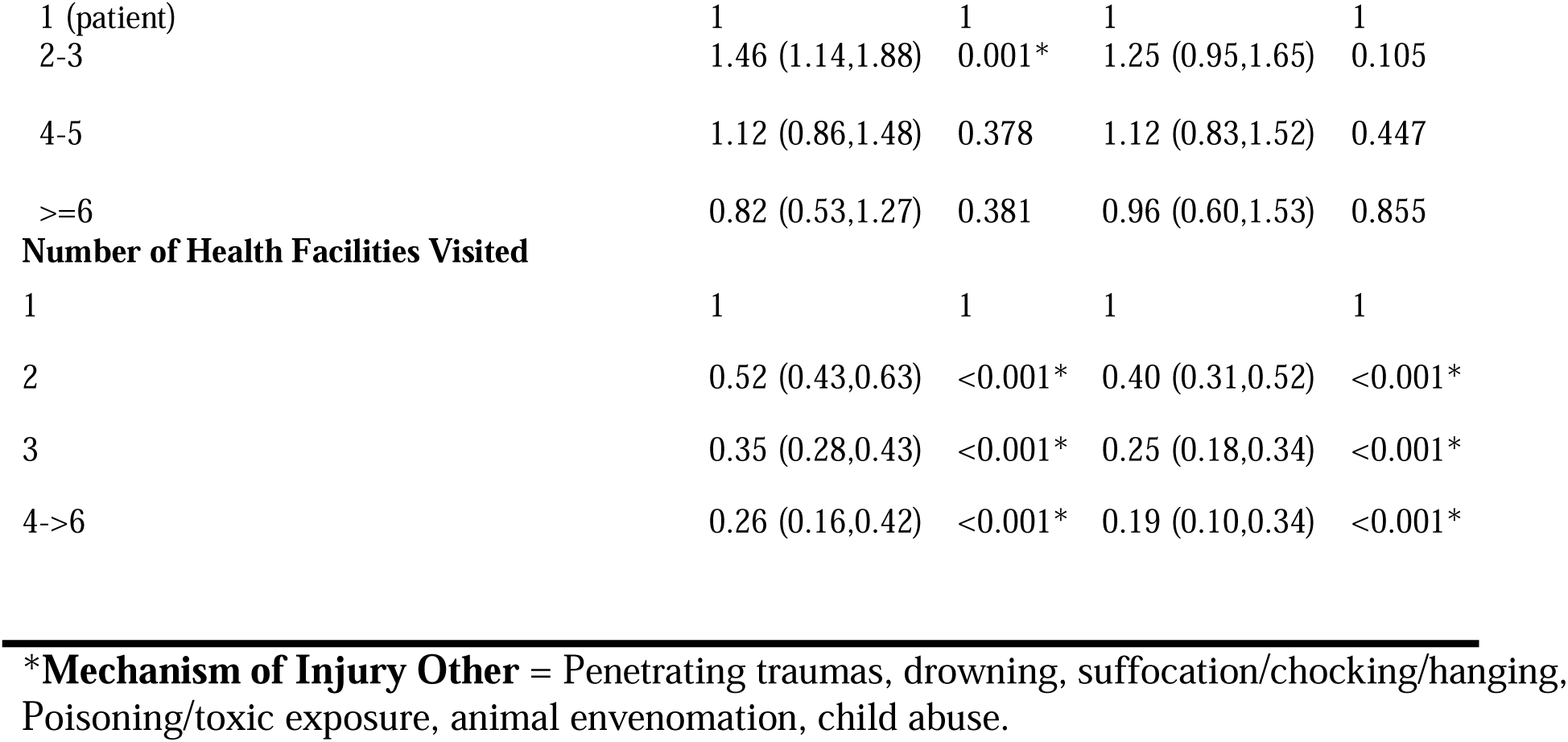
Cox proportional hazards regression showing crude and adjusted hazards ratios for time from injury to definitive care in hours (N=782).

| Variable | Crude HR |  | Adjusted HR |  |
| --- | --- | --- | --- | --- |
|  | HR (CI) | P-value | HR (CI) | P-value |
| **Age Group** |  |  |  |  |
| Infant (0-1 year) | 1 | 1 | 1 | 1 |
| Toddler (2-3 years) | 0.70 (0.51,0.97) | 0.032* | 0.70 (0.48,0.98) | 0.036* |
| Preschool (4-5 years) | 0.67 (0.49,0.91) | 0.011* | 0.68 (0.47,0.98) | 0.037* |
| Child (6-11 years) | 0.69 (0.52,0.92) | 0.015* | 0.60 (0.43,0.84) | 0.003* |
| Preteen (12-14 years) | 0.52 (0.37,0.74) | 0.001* | 0.71 (0.48,1.06) | 0.081 |
| Teen (15-17 years) | 0.65 (0.46,0.92) | 0.015* | 0.57 (0.38,0.84) | 0.005* |
| **Residency** |  |  |  |  |
| Moshi Urban | 1 | 1 | 1 | 1 |
| Moshi Rural | 0.78 (0.64,0.96) | 0.015* | 0.82 (0.67,1.01) | 0.059* |
| Hai | 1.32 (1.08,1.63) | 0.008* | 1.38 (1.11,1.71) | 0.004* |
| Arusha | 0.51 (0.36,0.73) | <0.001* | 1.01 (0.67,1.53) | 0.944 |
| Other | 0.50 (0.36,0.61) | <0.001* | 0.82 (0.60,1.14) | 0.241 |
| **Sex** |  |  |  |  |
| Male | 1 | 1 | 1 | 1 |
| Female | 1.02 (0.87,1.18) | 0.835 | 1.00 (0.85,1.18) | 0.997 |
| **Lives With** |  |  |  |  |
| Single parent | 1 | 1 | 1 | 1 |
| Both parents | 1.00 (0.78,1.29) | 0.985 | 0.99 (0.75,1.30) | 0.928 |
| Grandparent & Other | 0.66 (0.46,0.94) | 0.022* | 0.58 (0.40,0.87) | 0.008* |
| **Mechanism of Injury** |  |  |  |  |
| Road traffic injury | 1 | 1 | 1 | 1 |
| Blunt force/Stuck/Hit | 0.53 (0.41,0.69) | <0.001* | 0.51 (0.38,0.70) | <0.001* |
| Fall | 0.77 (0.61,0.95) | 0.016* | 0.43 (0.32,0.57) | <0.001* |
| Burn | 0.77 (0.59,1.00) | 0.050* | 0.47 (0.34,0.64) | <0.001* |
| Others | 0.77 (0.55,1.10) | 0.149 | 0.50 (0.33,0.74) | 0.001* |
| **Ambulance used** |  |  |  |  |
| No | 1 | 1 | 1 | 1 |
| Yes | 0.74 (0.61,0.88) | 0.012* | 1.43 (1.08,1.89) | 0.012* |
| **Payment method** |  |  |  |  |
| Insurance | 1 | 1 | 1 | 1 |
| Hospital exemption or child under 5 | 1.01 (0.62,1.63) | 0.970 | 0.53 (0.31,0.91) | 0.021* |
| None and abscond | 0.67 (0.29,1.53) | 0.340 | 1.08 (0.42,2.78) | 0.876 |
| Personal/family cash | 0.97 (0.76,1.25) | 0.825 | 0.72 (0.56,0.95) | 0.018* |
| Number of children living in the home, N (%) |  |  |  |  |
| 1 (patient) | 1 | 1 | 1 | 1 |
| 2-3 | 1.46 (1.14,1.88) | 0.001* | 1.25 (0.95,1.65) | 0.105 |
| 4-5 | 1.12 (0.86,1.48) | 0.378 | 1.12 (0.83,1.52) | 0.447 |
| >=6 | 0.82 (0.53,1.27) | 0.381 | 0.96 (0.60,1.53) | 0.855 |
| **Number of Health Facilities Visited** |  |  |  |  |
| 1 | 1 | 1 | 1 | 1 |
| 2 | 0.52 (0.43,0.63) | <0.001* | 0.40 (0.31,0.52) | <0.001* |
| 3 | 0.35 (0.28,0.43) | <0.001* | 0.25 (0.18,0.34) | <0.001* |
| 4->6 | 0.26 (0.16,0.42) | <0.001* | 0.19 (0.10,0.34) | <0.001* |

**\*Mechanism of Injury Other** = Penetrating traumas, drowning, suffocation/chocking/hanging, Poisoning/toxic exposure, animal envenomation, child abuse.

## Qualitative Results

Qualitative interviews with family caregivers and focus groups with healthcare providers revealed barriers to accessing definitive care and their drivers, that may contribute to the delays observed in the quantitative data. These barriers are organized by the Consolidated Framework for Implementation Research (CFIR) 2.0 domains: outer setting, inner setting, and characteristics of individuals (Figure 4, Table 3).(Damschroder et al. 2022)

**Figure 4.**
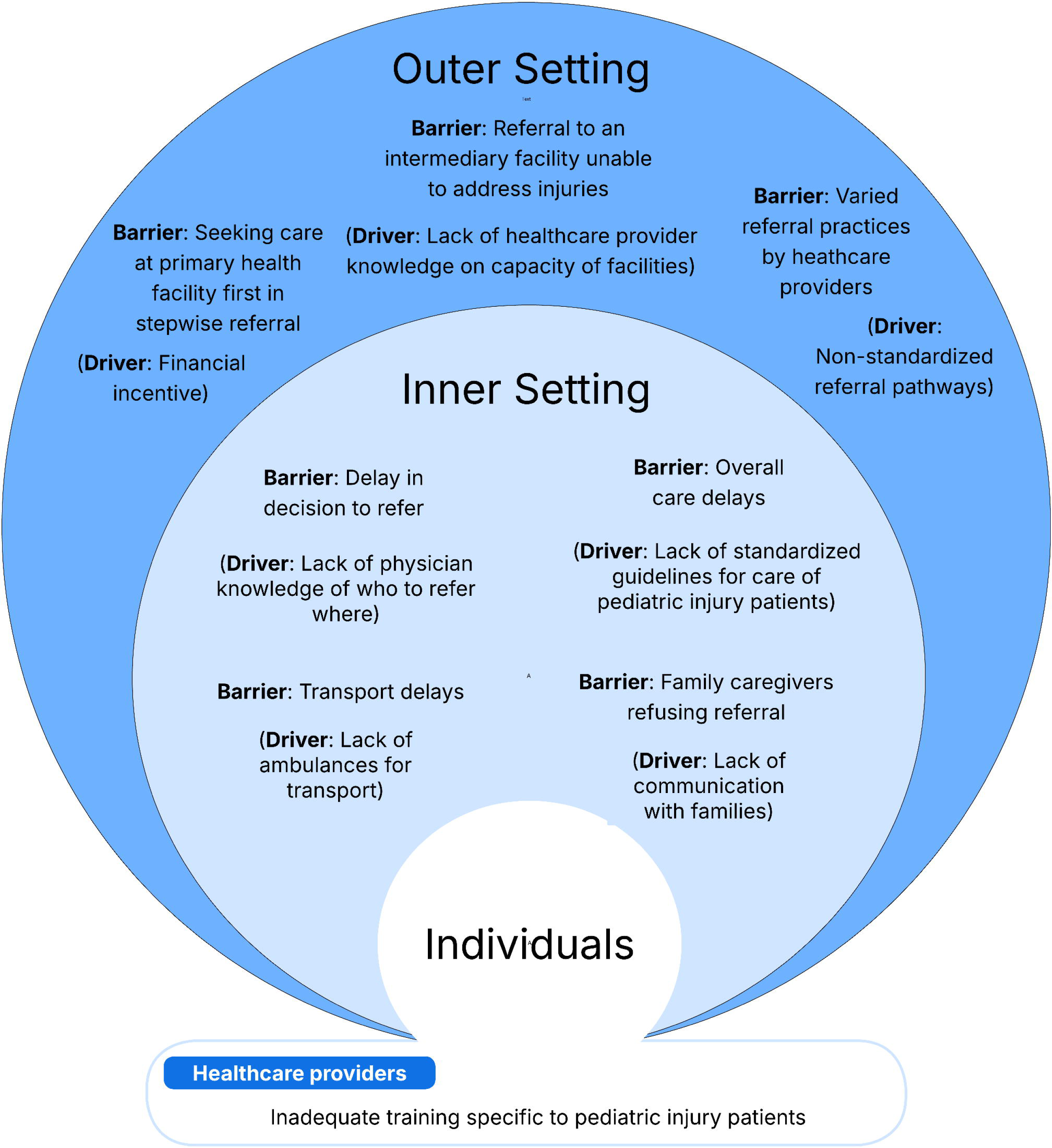
Barriers to accessing definitive care and their drivers, identified by family caregivers and healthcare providers. Barriers are categorized by the Consolidated Framework for Implementation Research (outer setting, inner setting, and characteristics of individuals) domains.(Damschroder et al. 2022) Alt text: Figure showing barriers to accessing definitive care and their drivers, identified by family caregivers and healthcare providers.

**Table 3.** Barriers to accessing definitive care, drivers of barriers, and representative quotes that lead to delays in referral of pediatric injury patients from the perspectives of family caregivers and healthcare providers.

| Barrier | Driver of barrier | Representative quote |
| --- | --- | --- |
| **Outer Setting** |  |  |
| Seeking care at primary health facility first (stepwise referral system) | Financial incentive – perception that care is discounted or free | “[Secondary Regional Hospital] is the designated regional referral hospital, and Pasua is a lower-level health center. According to the referral system, patients are first referred to the regional hospital before being sent to a national or tertiary hospital like [tertiary hospital name].” (IDI 028) |
| Referral to an intermediary facility unable to address injuries | Lack of healthcare provider knowledge on the capacity of health facilities at different levels | “I was received at the [secondary Regional Hospital] quickly, and the doctor advised me to go straight to [the third facility] with no treatment because my child’s eye condition was serious.” (IDI 029) |
| Varied referral practices by healthcare providers | Non-standardized referral pathways | “Sometimes we are unsure whether to refer to [tertiary hospital name] or manage the case here.” (FGD 04) |
| **Inner Setting** |  |  |
| Overall care delays | Lack of standardized guidelines for care of pediatric injury patients | “[We faced] delay made to transfer my injured child to [tertiary hospital name].” (IDI 027)<br>“There is a general guideline for treating pediatric children but no specific guideline for assessing pediatric injuries.” (FGD 02) |
| Transport delays | Lack of ambulances for transport | “I also waited for some time for the ambulance to come since it was not around, they had taken another patient to another hospital.” (IDI 001) |
| Delay in decision to refer | Lack of physician knowledge of who to refer where | “We spent almost one hour at [Outpatient Department] waiting for doctors to make decision [about referral].” (IDI 021) |
| Family caregivers refusing referral | Lack of communication with families | “They doctor just gave me a referral letter to go to [a secondary Regional Hospital], but they didn’t tell me why.” (IDI 027) |
| **Characteristics of Individuals** |  |  |
| Healthcare providers | Inadequate training specific to pediatric injury patients | “We need more training on assessing and stabilizing injured children.” (FGD 01) |

### Outer setting – the health system

Several features of the existing health system in Northern Tanzania create barriers for family caregivers and healthcare providers. Key themes included the stepwise referral system, insufficient capacity at intermediate facilities to treat injuries, and inconsistent referral processes.

#### Seeking care at the primary health facility first (stepwise referral system)

A few family caregivers mentioned the stepwise referral system when asked why they selected the first care site and why they were referred to a certain health facility for additional care. The current referral system in Northern Tanzania requires them to go to a primary-level health facility first and, when appropriate, be referred ‘up’ to the definitive care site. The explanation for this is that there is a financial incentive to doing this, as care is discounted when families present with a referral letter from the prior facility.

“If we had gone straight to [secondary Regional Hospital], they would have asked why we skipped the closest health center. We went to Pasua first so they could assess the child and refer us if necessary.” (IDI 028)

“The doctor said for moderate injuries the protocol allows them to start with [secondary Regional Hospital] first and if [secondary Regional Hospital] fails to take care of the injury they will transfer us themselves to KCMC.” (IDI 025)

#### Referral to an intermediary facility unable to address injuries

Many family caregivers described being referred from the first care site to an intermediary facility that could not address the injuries, and would rather have been referred directly to the tertiary facility. This unnecessary stop led to delays in treatment for these patients. This is explained by a lack of healthcare provider knowledge on the capacity of each facility.

“No, he didn’t get any treatment at [secondary Regional Hospital].” (IDI 029)

“At [the intermediary facility] there were a lot of delays, on arrival they also did an X-ray, applied a POP on the injured limb and said we should go back home and return on Tuesday because on Monday was public holiday. Bad enough when we return on Tuesday, they said they were not capable of managing my injured child so they need to transfer us to KCMC.” (IDI 021)

### Inconsistent referral processes

Healthcare providers reported a lack of standardized referral pathways that leads to inconsistent referral practices. They also described a lack of support for decision-making regarding injuries, which can lead to delays. Providers perceived this variability is because there is not a standardized referral pathway in the health system for pediatric injury patients.

“Right now, there are varied [referral] practices between staff members, and it can lead to confusion or delays. We need standardized guidelines for when and how to refer patients to KCMC, which would streamline the process.” (FGD 04)

### Inner setting – the health facilities that were the first sites for care

The inner setting was defined as decisions or care that occurred within health facilities that were the first site for injury care. Key themes included overall care delays at the first site, transport delays, delays in the decision to refer, and refusal of the referral by families, which was most often driven by a lack of clear communication by healthcare providers.

#### Overall care delays at first site of care

Many family caregivers described the biggest barriers being care delays at the first care site and the second care site. This is likely due to the lack of standardized guidelines for the care of pediatric injury patients.

“The big barrier was delays at Hai (first site) and [secondary Regional Hospital] (second site) to take care of my grandchild.” (IDI 018)

“There are no official or written protocols currently in use at Pasua Health Centre for the assessment of injured children. Most of the clinical decision making is based on Personal clinical judgment, team discussions during morning handovers, experience and occasional verbal instructions from senior staff.” (FGD 04)

#### Transport delays

Multiple family caregivers and healthcare providers described transport delays due to a lack of ambulances being a barrier to a timely referral. Even when the decision had been made to refer the patient and the family caregiver had accepted the referral, sometimes a patient had to wait many hours when an ambulance was out transporting another patient.

“We spent almost 2 hours at Pasua where most of the time we were waiting for the ambulance to come back because it had transferred another patient.” (IDI 026)

“Sometimes we face barrier of timely referral for a severely injured child when an ambulance has gone for another emergency.” (FGD 01)

#### Delay in the decision to refer

Family caregivers reported that triage of their injured child happened quickly, but it took more time to see the doctor and wait for a decision to be referred. Often patients were admitted to the primary health facility and the decision to refer was not made until the next day or the coming days. This is likely due to a lack of physician knowledge of who needs immediate referral. In addition, a few family caregivers described how referring providers were hesitant to refer to higher level of care even when requested by the family caregiver.

“It was on the 2nd day during ward round when one of the doctors told us they want to refer our injured child to KCMC due to severity of burn injury and also to receive precise care at burn unit in KCMC.” (IDI 013)

“After waiting for long time they then called us and said we need to give you referral to KCMC as we are not capable of managing her injury [here].” (IDI 018)

“On arrival at [facility] nothing was going on, even assessing the extent of burn injury was not done. Also, they put her on a dirty bed which might be source of infection, so I decide to request referral to another facility, but they were hesitating to do so.” (IDI 023)

#### Family caregivers refusing referral

Many family caregivers reported not understanding the reason for referral, which in some cases led to the families refusing or delaying the referral. This is likely due to them not understanding the reason for the referral, which is due to a lack of effective communication with families. Communication with families was a theme for the healthcare providers in the FGDs, as well.

“It was [the] second day after admission when one of the doctors told me they want to refer my child to KCMC for further care, but I disagreed and chose to come to [a secondary Regional Hospital].” (IDI 011)

“I had worry; why did they decide to transfer my injured child to higher facility?” (IDI 022)

“Sometimes [family] caregivers don’t understand the urgency and refuse or delay referral, especially if they worry about costs or distance.” (FGD 04)

### Characteristics of Individuals

Lastly, CFIR domains specific to the individuals were coded. Since the goal of this inquiry was to identify potential intervention avenues, characteristics specific to healthcare providers were coded for to inform solutions that account for the context (i.e., inner and outer settings) of Northern Tanzanian healthcare systems. Caregivers understand and seek care when an injury occurs. They often did not attempt first aid at home prior to seeking care. For burns, some home remedies were tried at home before seeking care. Healthcare providers indicated a lack of capacity and training as a fundamental barrier to providing appropriate and timely injury care.

“To be honest, most of us at Pasua Health Centre have not received any formal training specific to pediatric trauma care after our school education. What we apply now is mostly based on what we learned during medical school.” (FGD 04)

## Discussion

This study empoyed a multi-dimensional lense to investigate the health systems processes and barriers to rapid care for pediatric injury patients along their care pathways in Northern Tanzania. The purpose of this mixed-methods study was to use a patient registry to track time to definitive care and to examine systems-level barriers to efficient care from multiple perspectives. Among patients referred from first sites of care to definitive care, significant delays were observed that worsened with each referral to the next level of the health system. Qualitative data with both family caregivers and healthcare providers managing pediatric injury patients suggested various barriers that explain the delays at multiple levels, such as non-standardized referral pathways, a lack of knowledge on who to refer where, and inadequate training on care of pediatric injury patients. To improve the referral delays found in our study, a locally co-designed, multicomponent intervention need to be developed that gets patients more directly to definitive care, to decrease both the number of referrals each patient experiences and subsequent delays.

The delays observed in our study increased across health system segments. According to World Health Organization (WHO) standards, every child needing referral should be sent within 2 hours of arrival at the referring health facility.(2018) They should also receive appropriate pre-referral care, and the decision to refer should be made without delay.(2018) This 2-hour requirement is significantly lower than the median of each referral segment between health facilities in our study (range 7.6-140.8 hours). Thus, there is an opportunity to streamline the healthcare system by implementing an intervention at the first site of care to bring injured children to definitive care more quickly. Previous studies in Sub-Saharan Africa have reported similar delays, and these delays are associated with increased mortality risk.(Botchey et al. 2017; Boyd & Cowley 1983; Jin et al. 2021; Mock et al. 2009; Rogers et al. 2015; Sa’avu et al. 2014; Wisborg et al. 2011) Interestingly, many studies in SSA show that patients are often delayed in accessing care due to barriers such as geography, long travel times, financial constraints, transportation challenges, sparce ambulance services and unequal distribution of emergency services.(Ahmed et al. 2025; Pickering et al. 2022; Rolle et al. 2023; Sawe et al. 2022; Simon et al. 2013) However, in this study, the median time from injury to seeking first care was only 2.5 hours, and longer delays occurred after the patient was already within the health system. This argues for a need for an intervention at the level of the first interaction with the health system in Northern Tanzania.

After quantitatively demonstrating delays, family caregivers and healthcare providers were interviewed, who identified barriers and their drivers at the first site of care. Using CFIR, barriers were found at the level of the outer setting (health system), inner setting (first site of care), and individual (healthcare providers), including non-standardized referral pathways, a lack of knowledge on who to refer where, and inadequate training on care of pediatric injury patients, respectively. This demonstrates that the delays found are not due to a barrier at the level of one health facility, but rather a health systems-level problem that requires a health systems-level response with many stakeholders involved. This systems-based approach is unique in global injury. In the United States, Sasser et al. and the Centers for Disease Control used a health systems approach to implement the guidelines for field triage of injured patients with multi-level state and local community input.(Sasser et al. 2012) Local stakeholders co-designing a health systems intervention to solve challenges in their community can ensure that that intervention is locally and culturally relevant. Studies have demonstrated that stakeholder co-design can lead to innovative solutions and facilitate the adoption of interventions.(Al-Mondhiry et al. 2022; Baker et al. 2022; Brusco et al. 2022; McLachlan et al. 2022; Toomey et al. 2022) Thus, including the perspectives of family caregivers and clinical stakeholders at the first site of care will make this intervention more acceptable, feasible, and locally relevant.

The use of CFIR pre-implementation to guide the development of this intervention is supported by the CFIR literature in LMICs. A systematic review published in 2020 on the use of CFIR in LMICs included 34 studies, including 14 CFIR applications that guided data collection and 4 that framed/designed the intervention.(Means et al. 2020a) One study in Kenya used CFIR pre-implementation to develop a system-oriented intervention to improve service for children in district hospitals.(English 2013) Another study in Uganda used the CFIR pre-implementation to assess the implementation of national HIV policies on testing, treatment, and retention at health facilities.(McRobie et al. 2017) Two additional studies in maternal health in South Africa and public health in China also used CFIR pre-implementation to design the locally-relevant intervention.(Petersen Williams et al. 2015; Shi et al. 2016)

The barriers identified in this study at the multiple levels of the health system indicate a breakdown in the healthcare system’s referral pathway. To address this breakdown, a multicomponent intervention must be co-designed that more directly connects injured pediatric patients to definitive care. This could be addressed by creating a multicomponent, bundled intervention including standardized processes for assessing, caring for, and determining disposition for pediatric injury patients at the first site of care. Based on these findings, the necessary components should target healthcare providers at the first point of care, including training in pediatric injury care, pediatric injury guidelines, and referral decision support. Previous studies have shown that bundles of evidence-based interventions, when implemented consistently, result in better patient outcomes than when each is applied individually.(Gyawali et al. 2019; Sayed et al. 2026)

## Limitations

This study does have limitations. First, the findings are focused on two first care sites in Northern Tanzania, and these barriers may not generalize to other regions in the Tanzanian health system. However, the findings are relevant for this and similar regions in which local health centers and district hospitals are the first site of care for many injured patients. In addition, a limitation is that this study’s definition of definitive care assumes that the final place of care was the appropriate level for definitive care. However, it is possible that a child needed a higher level of care but did not get referred. The appropriateness of referrals or missed referrals will be assessed in future studies. Further, this study focused on pediatric injury patients and may not fully represent the experience of all emergency patients in this setting. Further work into non-injury emergency patients can be done to co-inform helath systems changes that have wider reaching impact. In contrast, a strength of this approach is that external researchers with close ties and connections to local care providers and systems were embedded in the team. The research was supported by local researchers who conducted interviews and assisted with data coding and interpretation. In short, the findings are locally relevant and potential solutions can be created that will be acceptable and feasible within this context.

## Conclusion

Every injured child who needs referral should be sent within two hours of arrival at the referring health facility according to current WHO standards. This study found health-system delays in referral of pediatric injury patients to definitive care in Northern Tanzania, particularly at the first point of care. Healthcare providers and family caregivers within the health system identified barriers at the outer setting (health system), inner setting (first sites of care), and individual level (healthcare providers). To reduce referral delays observed in this study, healthcare providers at the first site of care require a locally co-designed, multicomponent intervention that includes pediatric injury care training and referral decision support. Such an intervention would reduce delays to definitive care, thereby improving outcomes for pediatric injury patients in Northern Tanzania.

## Author Contributor Statement

Elizabeth M Keating contributed to the paper via the following: conception or design of the work, data collection, data analysis and interpretation, drafting the article, critical revision of the article, and final approval of the version to be submitted.

Rosalia Njau contributed to the paper via the following: data collection, data analysis and interpretation, critical revision of the article, and final approval of the version to be submitted. Joaquim Magoma contributed to the paper via the following: data interpretation, critical revision of the article, and final approval of the version to be submitted.

Baraka Moshi contributed to the paper via the following: data analysis and interpretation, critical revision of the article, and final approval of the version to be submitted.

Deusdedith Marandu contributed to the paper via the following: data collection, critical revision of the article, and final approval of the version to be submitted.

Emmanual Minja contributed to the paper via the following: conception or design of the work, data interpretation, critical revision of the article, and final approval of the version to be submitted.

Itikija Msuya contributed to the paper via the following: conception or design of the work, data interpretation, critical revision of the article, and final approval of the version to be submitted. Anord Anthony contributed to the paper via the following: data interpretation, critical revision of the article, and final approval of the version to be submitted.

Francis Sakita contributed to the paper via the following: conception or design of the work, data interpretation, critical revision of the article, and final approval of the version to be submitted. Matti Koenig contributed to the paper via the following: data analysis and interpretation, critical revision of the article, and final approval of the version to be submitted.

Edwin Shewiyo contributed to the paper via the following: data analysis and interpretation, critical revision of the article, and final approval of the version to be submitted.

Cassidy Huun contributed to the paper via the following: data analysis and interpretation, critical revision of the article, and final approval of the version to be submitted.

Sharla Rent contributed to the paper via the following: data analysis and interpretation, critical revision of the article, and final approval of the version to be submitted.

Getrude Nkini contributed to the paper via the following: data collection, critical revision of the article, and final approval of the version to be submitted.

Nancy Mmary contributed to the paper via the following: data collection, critical revision of the article, and final approval of the version to be submitted.

Heather T Keenan contributed to the paper via the following: conception or design of the work, data interpretation, critical revision of the article, and final approval of the version to be submitted.

Jennie L Hill contributed to the paper via the following: conception or design of the work, data interpretation, critical revision of the article, and final approval of the version to be submitted. Melissa H Watt contributed to the paper via the following: conception or design of the work, data interpretation, critical revision of the article, and final approval of the version to be submitted.

Catherine A Staton contributed to the paper via the following: conception or design of the work, data interpretation, critical revision of the article, and final approval of the version to be submitted.

Blandina T Mmbaga contributed to the paper via the following: conception or design of the work, data interpretation, critical revision of the article, and final approval of the version to be submitted.

Joao Ricardo Nickenig Vissoci contributed to the paper via the following: conception or design of the work, data analysis and interpretation, critical revision of the article, and final approval of the version to be submitted.

## Reflexivity Statement

The authors include 12 females and 9 males and span multiple levels of seniority, from research assistants to medical students to full professors in mentorship roles. The authors include 12 members from Tanzania and 9 members from the United States, two very different cultures. All authors have experience conducting research in Northern Tanzania.

## Ethical Approval

This study received ethical approval from the institutional review boards at the Tanzania National Institute for Medical Research (NIMR/HQ/R.8a/Vol.IX/4630; approved 5/21/2024), KCMC (Research Proposal No. 1450; approved 1/22/2024), and the University of Utah (IRB_00169039; approved 7/27/2023).

## Data Availability Statement

The data underlying this article cannot be shared publicly since the data for this manuscript are covered under a data sharing agreement. For data access requests, please contact our non-author KCMC representative Gwamaka William at.

